# Prevalence and Correlates of Parosmia and Phantosmia among Smell Disorders

**DOI:** 10.1101/2021.07.02.21259925

**Authors:** Robert Pellegrino, Joel D. Mainland, Christine E. Kelly, Jane K. Parker, Thomas Hummel

**Affiliations:** Monell Chemical Senses Center, Philadelphia, PA 19104, USA; Department of Neuroscience, University of Pennsylvania, Philadelphia, PA 19104, USA; AbScent, 14 London Road, Andover, Hampshire, UK; Department of Food and Nutritional Sciences, University of Reading, Whiteknights, Reading, UK; Smell & Taste Clinic, Dept. of Otorhinolaryngology, TU Dresden, Dresden, Germany

## Abstract

Among those many individuals who suffer from a reduced odor sensitivity (hyposmia/anosmia) some individuals also experience disorders that lead to odor distortion, such as parosmia (i.e., distorted odor with a known source), or odor phantoms (i.e., odor sensation without an odor source). We surveyed a large population with at least one olfactory disorder (N = 2031) and found that odor distortions were common (46%), with respondents reporting either parosmia (19%), phantosmia (11%), or both (16%). In comparison to respondents with hyposmia or anosmia, respondents with parosmia were more likely to be female, young, and suffered from post-viral olfactory loss (*p* < 0.001), while phantosmia occurred most frequently in middle-aged respondents (p < 0.01) and was more likely to be caused by head trauma than parosmia (*p* < 0.01). A higher prevalence of odor distortion was observed 3 months to a year after their olfactory symptom onset (*p* < 0.001), which coincides with the timeline of physiological recovery. Additionally, we observed that the frequency and duration of distortions negatively affects quality of life, with parosmia showing a higher range of severity than phantosmia (*p* < 0.001). Previous research often grouped these distortions together, but our results show that they have distinct patterns of demographics, medical history, and loss in quality of life.

## Introduction

Olfactory dysfunction affects a quarter of the population and with the advent of COVID-19 this number is likely to rise (Pellegrino, Cooper, et al., 2020). In addition to reduced odor sensitivity, individuals also experience odor distortion (Burges Watson et al., 2020; Keller & Malaspina, 2013; D. Leopold, 2002). Reduced sensitivity has been well described in the literature leading to better diagnosis and treatment (Hummel et al., 2017; Oleszkiewicz et al., 2019). Still, despite the differences between parosmia (i.e., distorted odor with a known source) and phantosmia (i.e., odor sensation without an odor source) (Hummel et al., 2017) most studies do not separate them. This is partly due to the large variance in their clinical presentation (Frasnelli et al., 2004) and because many patients report having both symptoms (Sjölund et al., 2017).

In general, when patients with parosmia inhale odorants their perception does not match their memory from before the distortion. In most cases of parosmia, the distorted odors are usually perceived as unpleasant (“cacosmia”), but there have been cases in which the distortions were pleasant (“euosmia”, (Landis et al., 2006)). Additionally, recent evidence suggest that specific odorants are more likely to trigger parosmia than others (Parker, Kelly, Smith, et al., 2021). Phantosmia, on the other hand, describes the perception of an odor in the absence of a source – there is only the illusion of a smell. Parosmia has been reported among 10% to 60% of olfactory dysfunction patients (Nordin et al., 1996; Parma et al., 2020; Reden et al., 2007) while the range is much smaller (3 – 16 %) for phantosmia (Bainbridge et al., 2018; Nordin et al., 1996; Ohayon, 2000; Rawal et al., 2016; Reden et al., 2007; Sjölund et al., 2017). These numbers indicate that incidences of parosmias and phantosmias are not rare, but the variance indicates that the reported frequency depends on the definition of parosmia or phantosmia.

Most parosmia appears to co-occur with olfactory loss due to viral infection, with the majority of cases resolving within a year (D. Liu T. et al., 2020; Nordin et al., 1996; Quint et al., 2001; Reden et al., 2007). Patients suffering from parosmia also had smaller olfactory bulbs compared to those with reduced sensitivity and no distortion (Mueller et al., 2005; Rombaux et al., 2009). In addition, parosmia was eliminated by preventing odors from entering the olfactory cleft in a case study (J. Liu et al., 2020). This supports a peripheral etiology and is consistent with the theory that parosmia results from mistargeting that occurs when olfactory sensory neurons regrow axons to the olfactory bulb during recovery (Holbrook et al., 2005; Hong et al., 2012a).

With phantosmia, peripheral origins of distortion may be maintained through abnormally active olfactory sensory neurons, loss of inhibitory neurons, or microbial infection creating a malodor (D. Leopold, 2002). The removal of the olfactory epithelium or even briefly occluding a nostril (irrelevant of side) has been shown to eliminate the olfactory illusions for some patients (D. A. Leopold et al., 1991, 2002). Many phantosmia patients have a history of head trauma (D. Leopold, 2002; Sjölund et al., 2017), psychiatric disorders (Croy et al., 2013; Frasnelli et al., 2004), and temporal lobe epilepsy as well as phantosmic episodes commonly preceding seizures, migraines and even weather in the form of auras (Aiello & Hirsch, 2013; D. Leopold, 2002), suggesting a central etiology from overactive neurons.

Patients with symptoms of olfactory distortion may suffer to a larger extent than those with a reduced sensitivity, as they are continually reminded of their problem. In fact, individuals with reduced perception of odors are often not even aware of their disorder (Oleszkiewicz et al., 2020; Oleszkiewicz & Hummel, 2019). However, most reports on odor distortions have not used a quantitative approach to compare them with anosmia and hyposmia– instead reporting quoted patient experiences. Here we compared them directly using a survey designed to gather information about parosmia and phantosmia. This quantitative approach allowed us to provide diagnostic criteria and reveal patterns of the disorder. Using this method, we saw several distinct differences among the disorders and created a severity metric for clinical use.

## Materials and Methods

### Sense of Smell Questionnaire

All data were collected through an online questionnaire that was created from prior research surveys (Frasnelli et al., 2004; Keller & Malaspina, 2013; Landis et al., 2010) and patient observations by the authors. It was designed to specifically address features of odor distortion (Supp. Appendix I). Two binary response (yes or no) questions accompanied by a descriptive caption were used to create four groups of smell impairment:

A. Parosmia - the experience of distorted smells which have an obvious source: Do you have parosmia (distorted sense of smell)?
B. Phantosmia - the experience of smells that have no obvious origin: Do you experience smells that are not present (phantosmia)?

Participants who only chose A or B were classified as Parosmic and Phantosmic respectively while those choosing both were considered both Parosmic/Phantosmic. All other smell impaired participants were considered Anosmic/Hyposmic. The questionnaire used a branching design in which questions specific to these disorders were only presented to those who responded with “Yes” to the quality disorder. The survey was distributed globally in English with English speaking countries (UK and USA) representing the largest proportions of respondents. The survey was launched in parallel with a new informational website about smell loss (www.abscent.org) which had two parts: an area with information that could be accessed by anyone, and a “member area” with a closed forum, access to the Sniff Smell Training app, and other more premium features. Access to the member area was given to anyone who completed the survey. Survey data was collected between May of 2019 and October of 2020. This procedure was conducted according to the Declaration of Helsinki for studies on human subjects and approved by the University of Tennessee IRB review for research involving human subjects (IRB # 19-05253-XM).

### Statistical Analysis

We used a unimodal analysis to look at differences across groups. We used chi-square analysis for categorical responses and analysis of variance (ANOVA) for continuous responses. Responses were bootstrapped to provide confidence intervals using the boot package (Davison & Hinkley, 1997). Here, we resampled (with replacement) the responses 1000 times to estimate error in all comparisons and visualizations.

To determine degree of severity, three questions were considered - each were asked within each odor distortion question block (answering “Yes” to Paromsia or Phantosmia) and had the same options. Below is an example of the questions for Phantosmia/parosmia.

A. How often do you experience smells that are not present (for phantosmia) or how often do you experience parosmia (distorted sense of smell)? (daily, once every week, once every month)
B. How long does a phantosmia/parosmia episode last? (seconds, minutes, hours, days)
C. How would you describe your phantosmia/parosmia? (mild, strong)

These questions together had a low intercorrelation coefficient (0.55). Questions A and C loaded onto the same principal component with question C explaining less variance (89% compared to 77%). Therefore, question C was dropped and A and B were summed to create a severity score of the disorder. Analysis was done with the psych package in R (Revelle, 2017).

Two open-ended text questions describing distortions of parosmia or phantomsia underwent text analysis. Sentences were cleaned and words were spell checked with hunspell using a large English dictionary (Ooms, 2020). Sentimental analysis, using the scentimentr package (Rinker, 2019), was done at the sentence-level across participants that provided sentences longer than the 1^st^ quartile length of all sentences (> 7 words) and density plots were used to provide a visual representation. Average sentiment and negative emotion count of each sentence were then used as predictors for degree of severity. Furthermore, sentences were broken down into one word nouns with SpacyR (Benoit & Matsuo, 2018). Summary tables of counts were constructed and visually represented in wordclouds with the size representing the frequency using ggwordcloud (Pennec & Slowikowski, 2018).

All analysis were done in R (version 4.3) and the code along with data can be found here: https://osf.io/5ebjt/

## Results

Only participants reporting a olfactory disorder, 18 years of age or over, and not born with the smell problem (congenital) were considered in the analysis (N = 2031). From this large population with an olfactory disorder, we report that odor distortions are common to smell impairment (46%) with individuals reporting either parosmia (19%), phantosmia (11%), or both (16%) (Figure 1a). Exploratory analysis revealed individuals reporting “both” types of odor distortion did not represent parosmia and phantosmia evenly (Supp Fig. 1). Due to this heterogeneity, we excluded this population from the rest of the analysis leaving three groups – Anosmic/Hyposmic, Parosmic, and Phantosmic. Parosmia and phantosmia showed distinct patterns, both from each other as well as from those with reduced sensitivity, in demographics, medical history, and impacts to quality of life. Using two questions, we were able to derive a severity score that influences many of these patterns.

**Figure 1.**
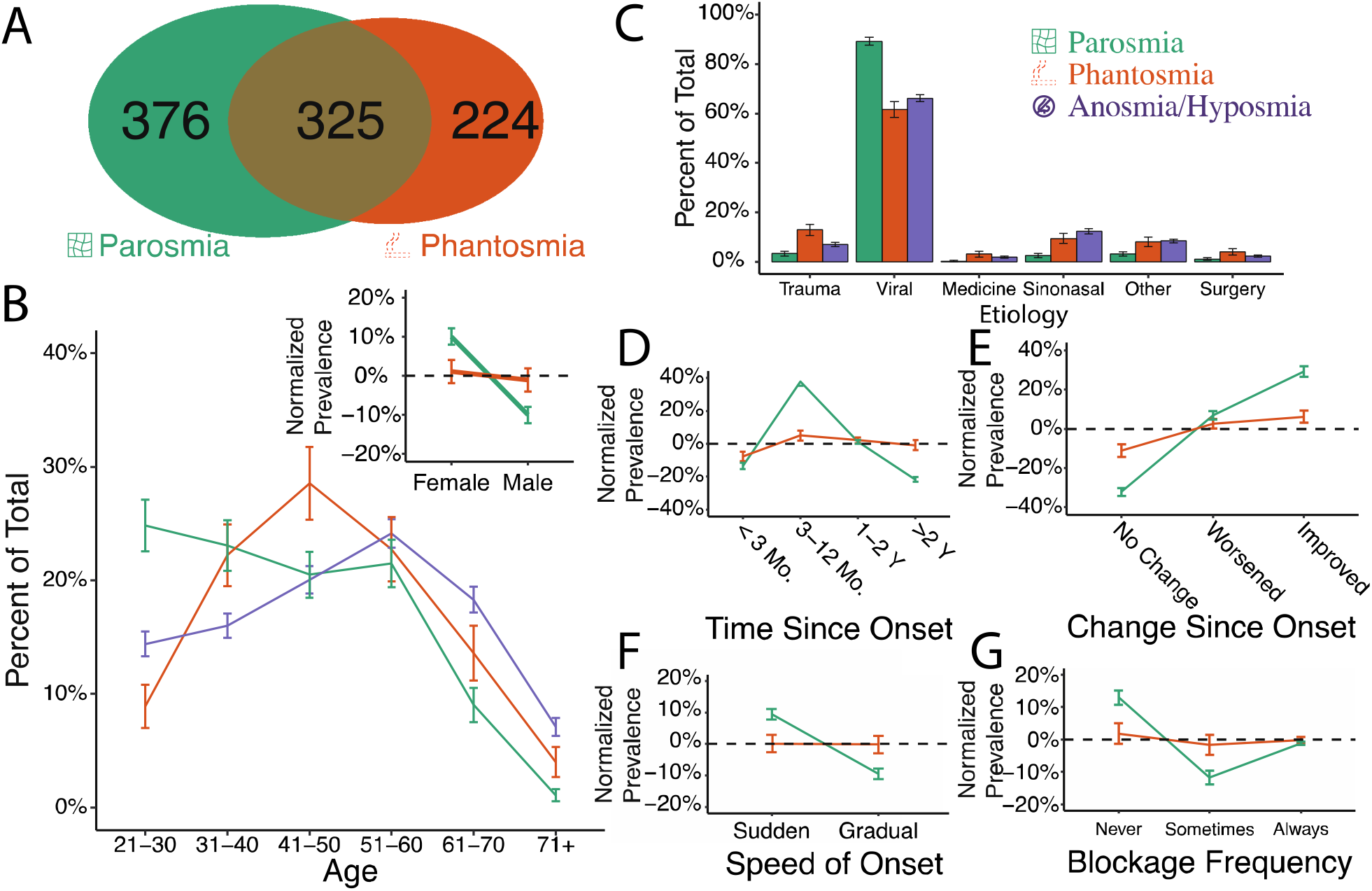
Parosmia, Phantosmia and Anosmia/Hyposmia are distinct disorders. (A) The number of study participants reporting having either parosmia, phantosmia or both. The three disorders were distinct in demographics (B), etiology (C), the time course of disease (D-F), and amount of congestion (G). Colors and icons represent olfactory disorders: green with a distorted grid icon represents individuals with parosmia, orange with an outlined cigarette icon represents individuals with phantosmia, and purple with a nose deny icon represents individuals with no parosmia nor phantosmia, but who reported an issue with smell (hyposmia/anosmia). Normalized prevalence represents the frequency difference between anosmia/hyposmia (baseline) and the other two olfactory disorders (parosmia or phantosmia). Error bars represent bootstrapped standard errors. Mo., Months; Y, Year

### Demographics and Medical History

Our sample was predominantly female (72%) with an age range from 21 to over 71 (see Supp Table 1). Respondents with parosmia were more likely to be female and younger than phantosmic (**χ**^2^ = 5.84, *p* = 0.047 and **χ**^2^ = 4.79, *p* < 0.001 respectively) or anosmic/hyposmic individuals (**χ**^2^= 14.12, *p* < 0.001 and **χ**^2^ = 4.62, *p* < 0.001 respectively) (Figure 1B). In contrast, phantosmia prevalence peaked for 41-50 year olds (**χ**^2^ = 2.82, *p* = 0.01) and anosmia/hyposmsia was more prominent in older individuals (61 and over; **χ**^2^ = 5.18, *p* < 0.001). There were no differences in gender between phantosmic vs. anosmic/hyposmic populations (**χ**^2^ = 0.08, *p* = 0.78).

The three most common etiologies resulting in an olfactory disorder are viral (70%), sinonasal disease (10%) and traumatic impact (8%) (Figure 1C). Among those with post-viral disorders, parosmia was the most ommon disorder (**χ**^2^ = 8.58, *p* < 0.001) and among those who suffered traumatic impact, phantosmia was the most common disorder (**χ**^2^ = 3.69, *p* = 0.006).

Compared to phantosmic and anosmic/hyposmic individuals, parosmia occurred suddenly (**χ**^2^ = 3.61, *p* < 0.001) with less nasal blockage (**χ**^2^ = 4.56, *p* < 0.001) (Figure 1F, G). Parosmia, compared to other olfactory conditions, was less likely to last more than two years (**χ**^2^ = 8.36, *p* < 0.001) and more likely to appear during recovery from the initial olfactory impairment (3 – 12 months) (**χ**^2^ = 13.35, *p* < 0.001) (Figure 1D). Similarly, parosmic individuals were more likely to say their condition was improving (**χ**^2 = 10.02, *p* < 0.001) and less likely to report their condition as unchanged (**χ**^2^ = 2.68, *p* = 0.02). Phantosmia, on the other hand, was more stable, with no change in improvement across time in comparison to the anosmic/hyposmic group (**χ**^2^ = 1.59, *p* = 0.33) (Figure 1E). Overall, parosmic individuals showed the most deviation from the other olfactory disorders (phantosmia and anosmia/hyposmia).

Parosmia is defined as distortion with an odor source, but the triggers for phantosmia are unknown. We report that all but one parosmic patient had specific sources that were distorted (99.7%, Figure 2A) while only a few phantosmic individuals had situations that triggered a distorted episode (17.0%). Sentences (N = 547) used to describe distortions for parosmia mostly had a negative sentiment, but there were positively described distortions (e.g., “my smell disorders are actually pleasant, flatulence smell like extra virgin olive oil and sometimes bubble gum”) (Figure 2B). Disgust was the highest emotion (Figure 2C, F(3) = 107.63, *p* < 0.001). Compiling words that trigger a distorted episode, parosmic individuals frequently reported foods that are roasted (coffee, meat) or contain sulfur (onion, egg, garlic). Phantosmic individuals instead reported places (room, house) or temporal events (e.g., time, week) while some referred to specific sensory (loud tv, cigarette smoke) or cognitive events (stress, memory).

**Figure 2.**
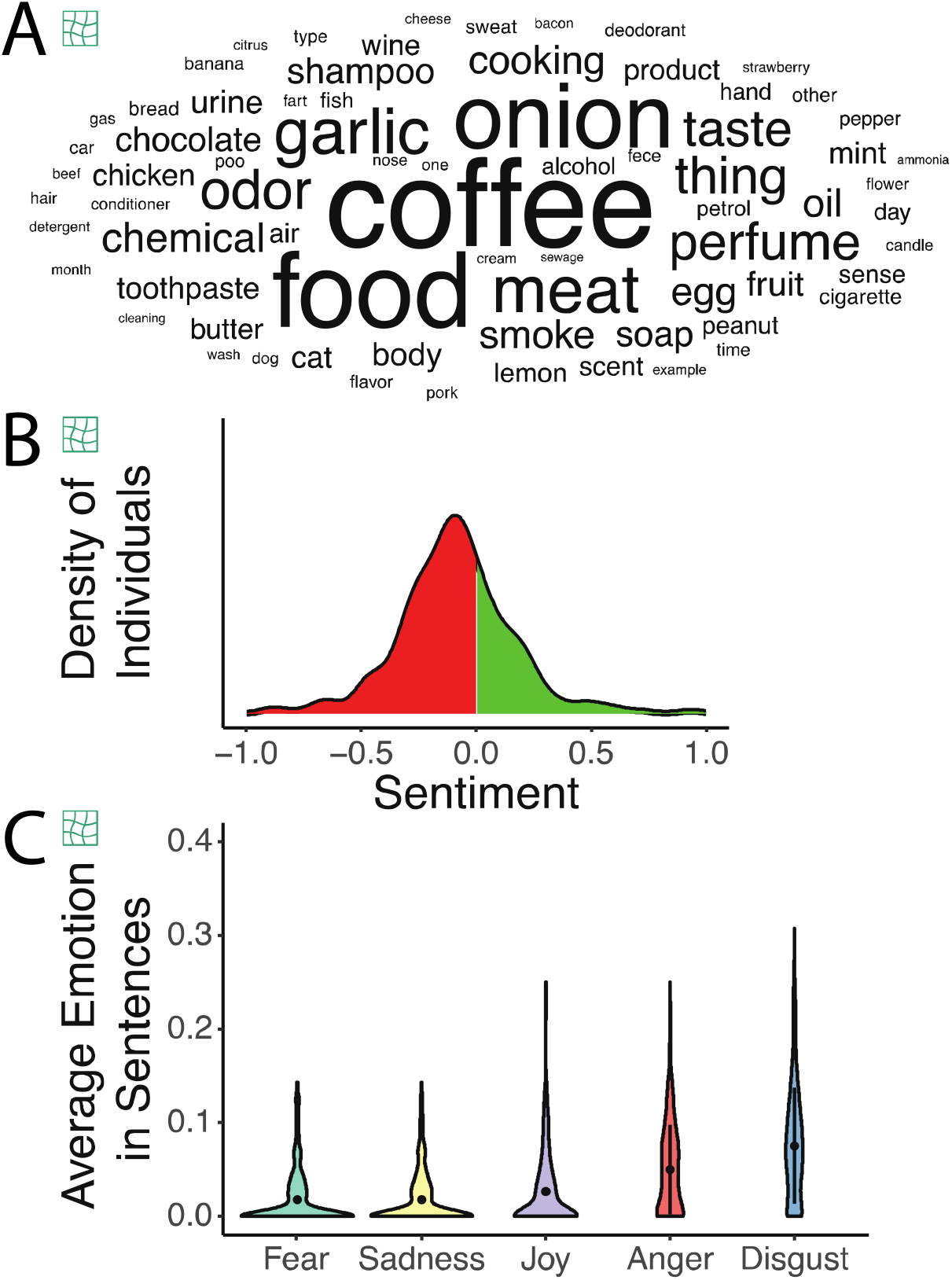
Text analysis of descriptions of parosmic episodes by individuals with parosmia. (A) Word cloud of nouns used to describe triggers of parosmia with size representing word frequency across 375 parosmics. (B) Distribution of sentences having a negative (in red) or positive (in green) sentiment. (C) Average emotions in sentences describing parosmia

### Quality of Life

All olfactory disorders affect overall quality of life, but each in different ways. Smell impaired individuals are concerned with failing to detect a hazard (> 50%) such as spoiled food (82.2%) followed by fire (72.8%) and gas (72.3%) (Figure 3A). Phantosmic and anosmic/hyposmic individuals showed a higher concern for failing to detect fire and gas than parosmic individuals.

**Figure 3.**
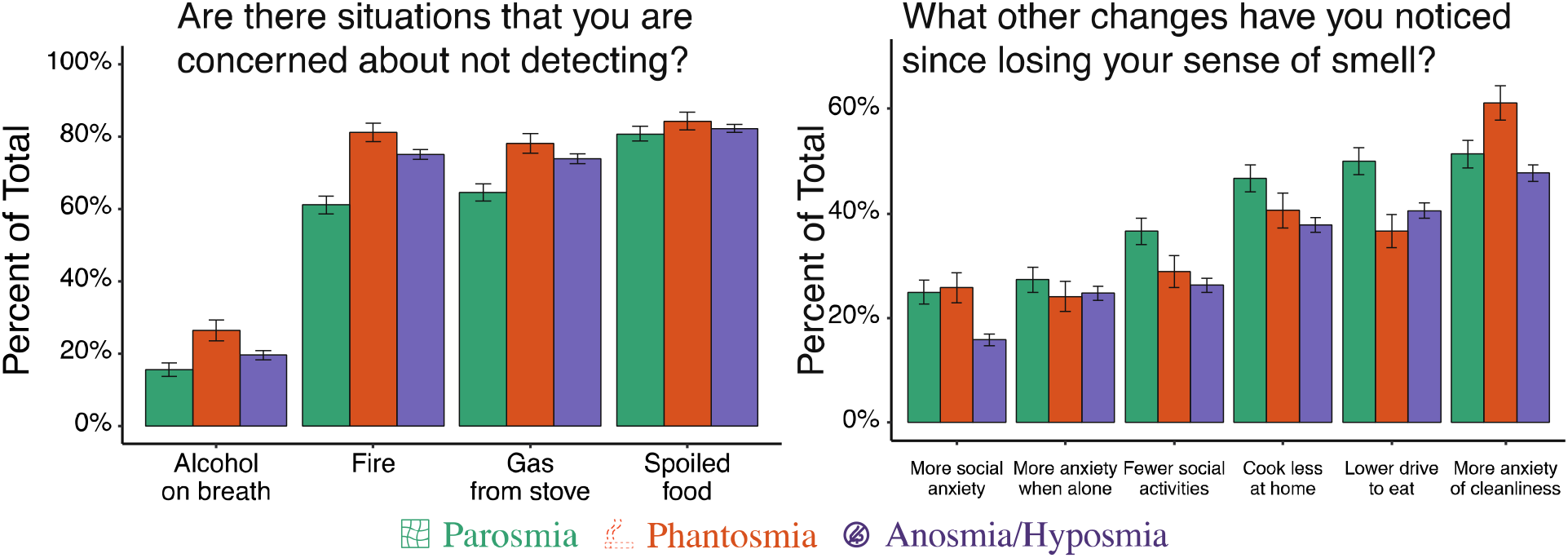
Impacts on quality of life. Percentage of respondents (A) concerned about failing to detect common hazards and (B) reporting changes in common behaviors. Error bars represent bootstrapped standard errors.

Other changes to quality of life include increased anxiety about being alone (25.2%), being in social settings (19.1%), cleanliness (50.4%), and cooking (40.2%) followed by a reported decrease in socializing (29.0%) and motivation to eat (42.1%) (Figure 3B). Among olfactory disorders, there was a higher anxiety for cleanliness among those with phantosmia and those with parosmia had a lower motivation to eat, cook and socialize. Both olfactory disorders reported more social anxiety than anosmic/hyposmic. Parosmic individuals also found it difficult to adjust to their disorder (**χ**^2 =3.76, p < 0.001) which might be a result of its acute nature during recovery. Phantosmics reported changes in their weight, with some gaining and others losing weight since the onset of the disorder (**χ**^2 = 5.27, p < 0.001) (Figure 4C). Intimacy was altered among 24% of respondents, but there were no differences across olfactory disorders (**χ**^2 = 5.40, *p* = 0.24).

**Figure 4.**
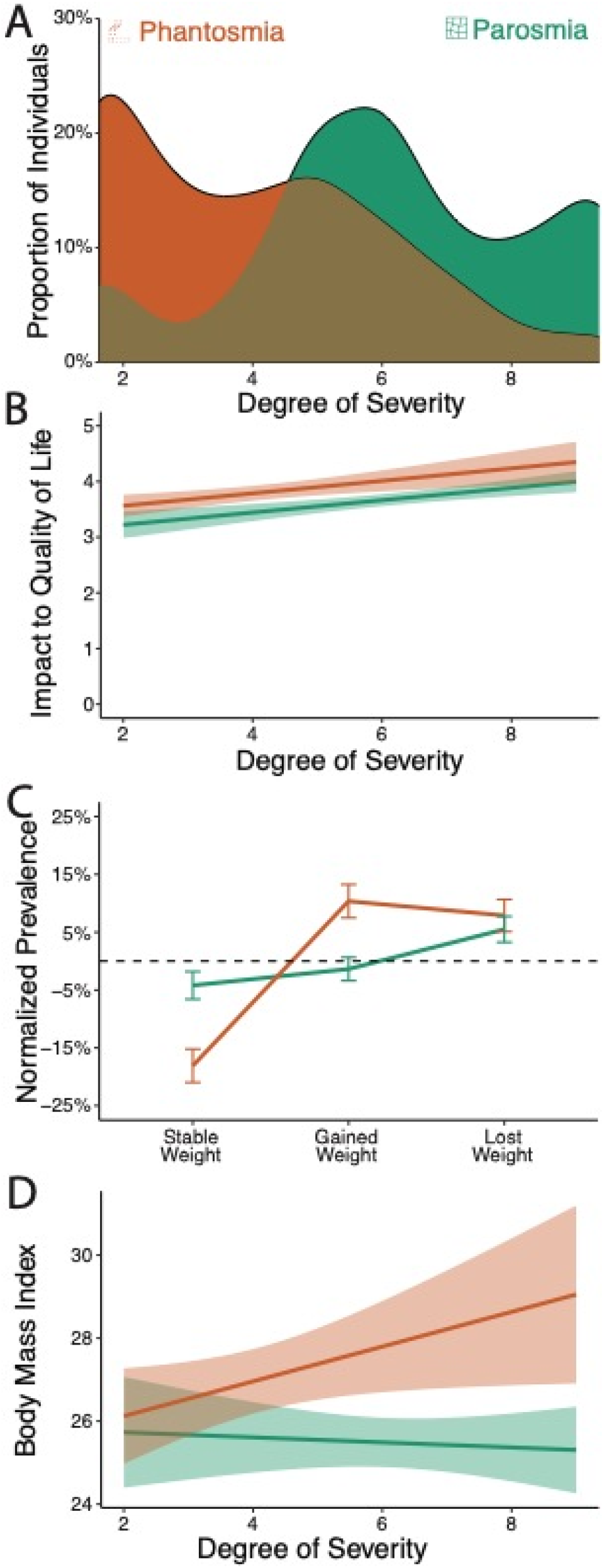
Degree of severity for parosmia and phantosmia. (A) Distribution of severity score among parosmic and phantosmic groups. (B) The severity score correlates with the reported impact of the olfactory disorder on their quality of life. Error bands represent 95% confidence intervals. (C) Differences in frequency of weight fluctuation. Error bars represent bootstrapped standard errors. (D) Relationship between degree of severity score and body mass index. Error bands represent 95% confidence intervals.

### Developing a Severity Score

A single scale of severity from structured questions has proven to be a clinically useful measure for parosmia, and here, we extend this idea to phantosmia (Landis et al., 2010). We combined the frequency and duration of distortion episodes to develop a severity score for both phantosmia and parosmia. Increases in the overall severity of the disorder affects the quality of life of individuals suffering from these disorders. Those with parosmia show a higher severity score than those with phantosmia (B = 1.96, t = 11.66, *p* < 0.001); Figure 4A), and an increased severity score was inversely correlated with overall quality of life for both disorders (B = -0.39, t = 4.16, *p* < 0.001; Figure 4B). More specifically, BMI was correlated with severity score for those with phantosmia, but not parosmia (b = 0.05, t = 1.79, *p* = 0.07; Figure 4C). As determined by the sentiment analysis, there was no relationship between severity score and negative emotions (B = -1.35, SE = 2.45, t = 0.55, *p* = 0.58), the type of emotion (F(3) = 0.02, *p* = .99), or overall sentiment (B = -0.53, SE = 0.49, t = 1.09, *p* = 0.28).

## Discussion

To date, little attention has been given to parosmia and phantosmia– with studies often lumping them together rather than studying them separately. Our study reveals some distinct differences between parosmia and phantosmia, as well as from hyposmia/anosmia. They are common olfactory impairments, with half of the participants with smell dysfunction reporting these disorders. Both parosmia and phantosmia vary in severity and are distinct in terms of demographics, medical history, and quality of life issues. Our survey also suggests that parosmia and phantosmia have distinct underlying mechanisms.

### Parosmia

Parosmia represents a distortion of smell when an odorous source is present. Instead of smells becoming weaker, as described in hyposmia/anosmia, they change in quality such that perceived smells are not the same as patients remember from before the onset of parosmia. In our survey, there is a distinct demographic that more commonly experiences parosmia – individuals who are younger, female, and recovering from a virus.

In general, there is a negative correlation between age and recovery from smell loss, such that losing smell at an older age results in slower recovery. One possibility is that parosmia is a symptom of recovery, and those who are older have a smaller chance of developing parosmia (Cavazzana et al., 2018; Hummel & Lötsch, 2010; London et al., 2008; Ogawa et al., 2020; Reden et al., 2006). Supporting this idea, individuals in the early stages of recovery from smell loss who report parosmia also reported more improvement over time than those with either phantosmia or a simple reduction in smell. Others have reported this co-occurrence of parosmia through times of recovery (D. Liu T. et al., 2020; Nordin et al., 1996; Quint et al., 2001; Reden et al., 2007). Its presence indicated faster return to the sense of smell in some studies (D. Liu T. et al., 2020; Reden et al., 2007), but not others (Hummel & Lötsch, 2010). This discrepancy may be due to patient age, since older patients have reduced olfactory regenerative capacity (Mobley et al., 2014).

Past research has reported parosmia commonly occurs with olfactory loss due to viral infection and frequently resolves within a year of the incident, with only 26% of an initial parosmic patient sample (N = 112) having parosmia after 14 months (D. Liu T. et al., 2020; Nordin et al., 1996; Quint et al., 2001; Reden et al., 2007). Similalry, in a study by Damm and coworkers (2014), 26% from a group of 47 initially parosmic patients reported no parosmia after an observation period of 4 months (Damm et al., 2014). Parosmia was the most prevalent outcome among post-viral disorders in our sample (nearly 90 % of parosmics) while parosmia had the lowest prevalence among those suffering from head trauma or conductive loss etiologies (e.g. polyps). As mentioned, patients with parosmia also showed higher prevalence of the disorder after the initial incident (> 3 months – 1 year), not during, and did not show issues with nasal patency.

Leading theories for parosmia suggest a peripheral origin of the disorder. Although these patients do show differences in neural activation (Iannilli et al., 2019), this might be a downstream effect. In fact, in hyposmic patients with parosmias, olfactory bulb volumes have been shown to be smaller compared to hyposmic patients without parosmia (Mueller et al., 2005; Rombaux et al., 2009). In neurogenesis, the axons of newly born sensory neurons must find the proper targets in the olfactory bulb. Abnormalities may occur during the process (Murai et al., 2016; Schwob et al., 2017), such that a sensory neuron tuned to one odor mistakenly stimulates an area of the bulb that signals the presence of a different odor. Axons reach the bulb approximately 1-3 months after injury, which matches the timing of parosmia in this survey. Taken together, our data support a peripheral cause of distortion that may result from a variety of mechanisms related to recovery such as differences across olfactory sensory neurons in time to recover or a mismatch in rewiring in the olfactory bulb. This is supported by animal models where olfactory maps significantly change after regeneration of ablated neurons, leading animals to have to relearn the meaning of odors (Yee & Costanzo, 1998) and this is most likely due to mistargeting by a receptor-defined subset of peripheral neurons (Christensen et al., 2001; Holbrook et al., 2005).

Parosmic patients showed higher disturbances to their social life, leading to an avoidance of social and eating activities. In comparison to hyposmia/ansomia, this did not lead to any associated behavioral outcomes that we measured, such as weight fluctuation, but more rigorous assessments are warranted (Mattes & Cowart, 1994). For instance, we clearly show that individuals with parosmia are reminded of their disorder regularly, which has been hypothesized as a reason for greater disruption in daily life (Croy et al., 2013; Frasnelli & Hummel, 2005; Hong et al., 2012b). These patients also report more difficulty adjusting to their disorder, which may explain a recent report showing higher depression and anxiety symptoms in this patient group (Lecuyer Giguere et al., 2020).

The distortions experienced describe a common thread of sources (e.g. coffee) that has been reported in the literature and there is little doubt that the terms used to describe these distortions generally have a negative valence associated with them (dirty, sewage, unpleasant, rotting, disgusting, sickly sweet and vomit-inducing) (Burges Watson et al., 2020; Keller & Malaspina, 2013; Parker, Kelly, Smith, et al., 2021). Although of our question about distortions had a negative phrasing, “Which odors do you find particularly unpleasant and distorted? (Describe in as much detail as possible)”, individuals still reported some positive changes. Looking at the positive and negative sentimental sentences, there seems to be a valence shift in which odors commonly perceived as positive are described negatively, but a few, usually related to body odors, shift from negative to positive. For instance, fecal smells may turn pleasant whereas coffee becomes unpleasant. One explanation for this shift from negative to positive is that some of the key aroma compounds responsible for the strong and usually repulsive smell of feces were not perceived at all by those with parosmia (Parker, Kelly, & Gane, 2021). In the absence of these potent odors, other pleasant compounds may dominate perception of the mixture.

### Phanstosmia

Phantosmia is an odorous experience when there is no odor source present. These odor phantoms may be high or low in intensity and may be familiar or unfamiliar odors, but others nearby do not perceive the smell. Unlike previous reports done at a population level (Bainbridge et al., 2018; Sjölund et al., 2017), in our sample females were not more prone to phantosmia (p = 0.78). This difference in findings may be due to previous studies categorizing parosmia and phantosmia together. For instance, a population level study found females to be almost twice as likely to have phantosmia than men, but the group under study also reported they were 6 times more likely to have parosmia thus representing a heterogeneous group (Sjölund et al., 2017). However, our results do agree with previous findings regarding age, in which individuals between 40 and 60 years of age were more likely to have phantosmia than older individuals (> 60 years) (Bainbridge et al., 2018).

Phantosmia was the most common olfactory disorder to have suffered a head trauma. Phantosmic patients have previously been reported to have a history of head trauma (D. Leopold, 2002; Sjölund et al., 2017), as well as psychiatric disorders (Croy et al., 2013; Frasnelli et al., 2004), temporal lobe epilepsy and phantosmic episodes commonly preceding seizures, and migraines in the form of auras (Aiello & Hirsch, 2013; D. Leopold, 2002). Additionally, we show that phantosmic patients had more sinonasal diseases (e.g. polyps) and more blockage than those suffering from parosmia. This suggests that at least some of these phantom odors do not come from an odorous source, as airflow is needed to carry volatiles.

Mechanisms of phantosmia are largely unknown, although theories of peripheral and central etiologies have been proposed. According to the central origin theory, one possibility for experiencing phantoms may be irregular firing of neurons in the primary and secondary olfactory processing areas in the brain. Input from the olfactory bulb excites cells in the piriform cortex to induce odor perception; however, this includes only a fraction of cells in that area with the majority not directly stimulated by OB input, but modulated by a recurrent network (Franks et al., 2011). Piriform cortex cells are additionally dampened by inhibitory neurons across the allocortex which modulate odor-related activity (Bolding & Franks, 2018). Recently, a paper showed excitation in this area limits inhibitory neurons leading to possible epileptogenic behavior (Ryu et al., 2021). In fact, traumatic brain injury patients with smell loss typically have an intact piriform that responds to odors at the same rate as those without impairment (Pellegrino et al., 2021). Hallucinations may also elicited by stimulation to the olfactory bulb and tract (Kumar et al., 2012). Therefore, increased excitation from irregular activity of damaged brain structures along with possible downplaying of inhibitory neurons may lead to increased phantom episodes. Similarly, hallucinations in other senses can be due to overactive neurons in the epithelium or in the brain (), and removal of the olfactory bulb () or olfactory epithelium (Leopold et al., 1991, 2002) decreases phantoms.

Phantosmic patients reported worries about not being able to detect hazards (fire, gas) that might be noticed through smell. Only a few (∼20%) had recurring situations that triggered a phantom episode, describing these triggers as place, temporal, or cognitive events. Additionally, phantosmic patients reported more changes in weight, with individuals experiencing more severe phantoms having an increase in weight (measured by BMI). Fluctuation in appetite with olfactory dysfunction occurs due its involvement in metabolic status (Guzmán-Ruiz et al., 2021) and this sometimes leads to changes in food preference (Pellegrino, et al., 2020) and weight (Kershaw & Mattes, 2018). However, it is difficult to say whether phantoms are causal. For example insulin-dependent diabetics, who often have a comorbidity of being overweight, were twice as likely to experience phantom odors (Chan et al., 2018). This may warrant additional studies to replicate our findings and delve into specific dietary changes and whether adiposity is related to a higher rate of smell phantoms.

### Study Limitations

Our study is based on cross-sectional data from a survey, therefore, direction of associations among variables with time cannot be established and this may undermine our causal inference in recovery for parosmia patients. Longitudinal studies with this patient group should be done to confirm our results. There is also an issue of subjective reporting for olfactory disorder types. We provided clear definitions of each type of distorted disorder, but it is difficult to exclude the possibility that some participants did not understand the meaning of parosmia and phantosmia. Indeed, we have a large category of respondents reporting both parosmia and phantosmia that were not included in the analysis as it did not fall into a separate group and were difficult to interpret. The group who reported both parosmia and phantosmia could be either those who experience distortions from both known and unknown sources, or those with a hybrid scenario where they recognize the source of their distortions, but for whom the distorted smell persists for hours or days after the stimulus has disappeared. Patients have reported this before as a “smell lock” and clinicians have referred to it as olfactory perseveration (Parker et al. 2021a). Lastly, we sampled from a smell loss group that has interest in their disorder (actively joining an interest group / charity) and there was overlap in our sampling times with the COVID-19 pandemic. Post-viral loss was the most prominent etiology in our sample – this might be due to our sampling times overlapping with the COVID-19 pandemic, where smell loss is a prominent symptom of the disease. Thus, our results may not represent the typical patient population at large. We report half the smell loss patients to experience odor distortion and this should be considered a liberal estimate.

## Conclusion

Two common symptoms of olfactory dysfunction, parosmia and phantosmia, represent distinct conditions that, along with hyposmia and anosmia, have characteristic patterns of medical history, demographics and how they affect quality of life. They are not rare, with almost half our sample reporting symptoms, and cause additional distress typically after an initial olfactory dysfunction starts to resolve. The mechanisms for distinct features of these smell distortions should undergo consideration in the clinic and research setting. If parosmia relates to neurogenesis, what does the character of distortion tell us about the underlying population of recovery neurons? Similarly, if phantosmia is centrally caused what does this tell us about our perception of reality? Distortions among olfactory disorders may provide answers to interesting research questions.

## Data Availability

All analysis along with data can be found here: https://osf.io/5ebjt/

## Notes

Conflict of Interest: The authors declare no conflict of interest.

### Competing Interest Statement

The authors have declared no competing interest.

### Funding Statement

No external funding

### Author Declarations

This procedure was conducted according to the Declaration of Helsinki for studies on human subjects and approved by the University of Tennessee IRB review for research involving human subjects.

